# Fairness in infectious disease modeling

**DOI:** 10.1101/2025.11.11.25339990

**Authors:** Yuhan Li, Nicolò Gozzi, Nicola Perra, Michele Tizzoni

**Affiliations:** School of Mathematical Sciences, Queen Mary University of London, London, UK; ISI Foundation, Turin, Italy; The Alan Turing Institute, London, UK; Department of Sociology and Social Research, University of Trento, Italy

## Abstract

The concept of fairness has been extensively examined within the domains of Machine Learning and Artificial Intelligence more broadly. It remains, however, largely underexplored in the field of Computational Epidemiology. Considering the substantial influence that epidemic models exert on public health policy, particularly in the context of outbreak preparedness and response, this shortcoming is of great relevance. Here, we propose a mathematical framework, grounded in core principles from Social Epidemiology, for evaluating the fairness of computational epidemic models. We begin by applying our framework to a range of epidemic modeling approaches and simulation scenarios, such as the initial spread of COVID-19 in London, New York, and Santiago de Chile, as well as the 2016 Zika virus outbreak in Colombia, demonstrating its consistent capacity to assess model fairness across diverse disease dynamics. Subsequently, we illustrate how our definition of fairness can be incorporated into the design of immunization strategies as a way to enhance health equity while simultaneously improving the overall effectiveness of such interventions. Overall, our results offer a new systematic methodology for quantifying fairness in computational epidemiology.

## 1 Introduction

Infectious disease modeling is a key tool for informing the response to outbreaks and guiding policymakers in the adoption of interventions^1^. During the COVID-19 pandemic, for example, computational models have been widely used to assess the impact of non-pharmaceutical interventions (NPIs), evaluate future scenarios, and design mitigation strategies^2^. Due to a limited understanding of the interaction between the dynamics of the epidemic and the social determinants of health, in several cases, such intervention strategies have amplified disparities rather than reduced them^3–6^. Indeed, the COVID-19 pandemic has been a stark reminder that infectious diseases do not affect populations equally^5,7^. Outbreaks are often characterized by massive inequities, resulting in disproportionate health impacts among vulnerable groups and minorities^8,9^.

Most of the epidemiological modeling frameworks available are unequipped to adequately capture the structural drivers of unequal health outcomes. Indeed, recent studies have highlighted how computational epidemiology lacks standard frameworks to address inequalities and health disparities systematically^10–12^. The urgent need for equitable models has led to a widespread call for epidemic models that can integrate aspects of equity into their mechanisms, leading to realistic descriptions of the epidemic burden among different socioeconomic and demographic groups^10,12–14^.

The lack of equitable computational modeling frameworks to simulate infectious disease epidemics is accompanied by a lack of standard methods to define and evaluate a model’s *fairness*. On the contrary, due to their increasing relevance in real-world decision making, the concept of algorithmic fairness has become central to Machine Learning and Artificial Intelligence^15,16^. Epidemic modeling has a relevant impact on policy decisions, but a commonly agreed definition of the fairness of infectious disease models remains elusive^17^. Previous studies have addressed the issue of fairness in infectious disease forecasting^18–20^. However, they mainly focused on racial/ethnic disparities associated with the burden of COVID-19 in the United States, leaving the search for a broader definition of fairness, applicable to different diseases and contexts, largely unexplored.

Here, we address this gap by proposing the first general definition of fairness for models of infectious disease spread, grounded in the theory of Social Epidemiology^21,22^. We define the fairness of an epidemic model as its ability to capture the (un)equal distribution of disease burden according to one or more equity-related features. Given a demographic or socioeconomic trait that is a known determinant of health and the disease dynamics under study, an epidemic model will be fair if it captures the observed inequality in health outcomes according to such a variable. We translate our definition into practice by providing quantitative fairness measures for different epidemic scenarios, introducing a set of metrics that quantify the fairness of a model in various cases. Following the traditional literature of Social Epidemiology^21^, we consider health inequalities in: (i) ordered social groups, (ii) unordered social groups, and (iii) spatially structured populations. In studying social inequalities, ordered social groups are those that exist along a clear hierarchical continuum—such as income levels—where members can be ranked from higher to lower status. Inequality in these groups is often analyzed in terms of relative positions and gradients of advantage or disadvantage. In contrast, unordered social groups—such as those based on gender, ethnicity, or religion—do not form a ranked continuum but are instead categorical distinctions. Inequality arises here from differential treatment, discrimination, or exclusion between groups, rather than from a measurable position along a social scale. As we will discuss, spatially structured populations can be considered a third case in which ordering may or may not be present.

For each type of social inequality, we introduce a specific metric that quantifies a model’s fairness: the relative concentration index (RCI) fairness, the Theil index fairness, and spatial fairness. The first quantifies the accuracy of the model in capturing the health gradients associated with naturally ordered social groups, such as income, education, or age. The second score assesses the fairness of the model in the case of unordered social groups, such as racial or ethnic groups. The third measure assesses a model’s fairness in capturing the geographic disparities of disease burden, regardless of whether this is associated with ordered or unordered groups.

We first explore the application of our definitions of fairness through numerical simulations of the initial spread of COVID-19 in the United States, the United Kingdom, and Chile, as well as the Zika virus in Colombia. The results show that our definitions of fairness can be consistently applied to a range of modeling approaches and disease scenarios. Second, we demonstrate how the proposed fairness scoring methods could inform policy responses by enhancing the equity of decisions in an epidemic outbreak. Indeed, by simulating a proof-of-concept vaccination scenario, we demonstrate that our fairness metrics not only represent a novel equity-aware subgroup model validation tool but can also provide guidance for a more equitable allocation of resources across groups.

## 2 Results

### 2.1 Fairness of infectious disease models and health inequalities

Mathematical and computational models of infectious diseases usually incorporate a representation of the population of interest. It is common to assume that some population traits are more relevant than others for disease transmission and health outcomes. The simplest models, like the well-known SIR model, assume individuals in a population to be indistinguishable apart from their health status (i.e., susceptible, infected, or recovered)^23^. As models grow in complexity, they can integrate more traits to capture the characteristics that are deemed important for explaining disease outcomes. A classic example is the introduction of age groups and mixing patterns as a way to capture the observed heterogeneities in contact rates and health outcomes by age^24^. We generally refer to socioeconomic or demographic characteristics as equity-related since the choice to introduce one or more of them into a model translates into the model’s ability to capture the health inequalities associated with them. In Social Epidemiology, health inequalities are generally described as differences wherein socially disadvantaged groups, those who have consistently faced marginalization or discrimination, systematically experience poorer health outcomes or higher health risks compared to more advantaged populations^22^. The decision about which equity-related characteristics to include should be based on data on which important health or social heterogeneities exist by group or on which groups are of specific interest to policymakers and public health officials. In particular, the choice should prioritize those inequalities that can be addressed through intervention by policymakers. Based on the concept of equity-related model features, we propose a general definition of fairness for epidemic models as follows.

We define the fairness of an epidemic model as the model’s ability to capture the (un)equal distribution of disease-related health outcomes according to one or more equity-related features. Given any demographic or socioeconomic variable that describes the population under study, an epidemic model will be *fair* if it accurately captures the observed distribution of health outcomes according to such variables.

The above definition of fairness incorporates the principle of *equal performance* from the machine learning domain^25^, requiring the maximization of a model’s accuracy for all groups in the population. However, it is important to note that, unlike machine learning algorithms, infectious disease models do not generate outcomes or predictions at the individual level, but often only at the population level. This implies that many popular fairness constraints adopted in machine learning, such as Demographic Parity, Equalized Odds, and Equal Opportunity^26^, cannot be easily translated to the context of epidemic models. In the following, we apply our definition by providing quantitative measures of the fairness of a model for different diseases, epidemic scenarios, and modeling assumptions. We follow a classic distinction in Social Epidemiology^21^, considering three different types of health inequality that correspond—in our framework—to three different measures of fairness. In particular, we consider the case of i) ordered social groups, ii) unordered social groups, and iii) spatially structured populations. This distinction is not merely conceptual but has direct practical implications, since measures of inequality and fairness developed

for ordered social groups typically cannot be applied to unordered groups. In all three cases, our approach to measuring the model fairness consists of three steps:

1. The identification of a measure of inequality to describe the health disparities observed in real data;
2. The characterization of the health disparities observed in a model output using such a measure of inequality;
3. The definition and application of a fairness metric that compares a model’s results with the real data according to the chosen measure of inequality.

Finally, we demonstrate how the proposed approach to quantifying fairness can inform policymaking by increasing the equity of health outcomes in public health interventions, such as vaccination campaigns.

### 2.2 Fairness in ordered social groups

We first consider the case of health inequalities associated with *ordered social groups*. Some demographic or socioeconomic traits imply an inherent ordering of the population subgroups, regardless of the health status of their members. For example, any trait measuring dimensions of the socioeconomic position of individuals implies a natural ordering, such as education, income, and wealth. Similarly, individuals’ age naturally defines ordered groups within a population. In the case of ordered social groups, a widely used measure of health inequality is the Relative Concentration Index (RCI)^27,28^. The RCI is derived from a health concentration curve, where the population is ordered by social group status, and the cumulative fraction of the population is plotted against their share of total ill-health. Fig. 1a illustrates the relationship between the RCI and the health concentration curve (right) for a fictitious disease, with death rates varying by income deciles of the population (left). The relationship between the RCI and the health concentration curve is similar to the relationship between the Gini coefficient and the Lorenz curve, with the difference that in a Lorenz curve, the population share is ranked by socioeconomic variables on both axes^29^.

**Figure 1.**
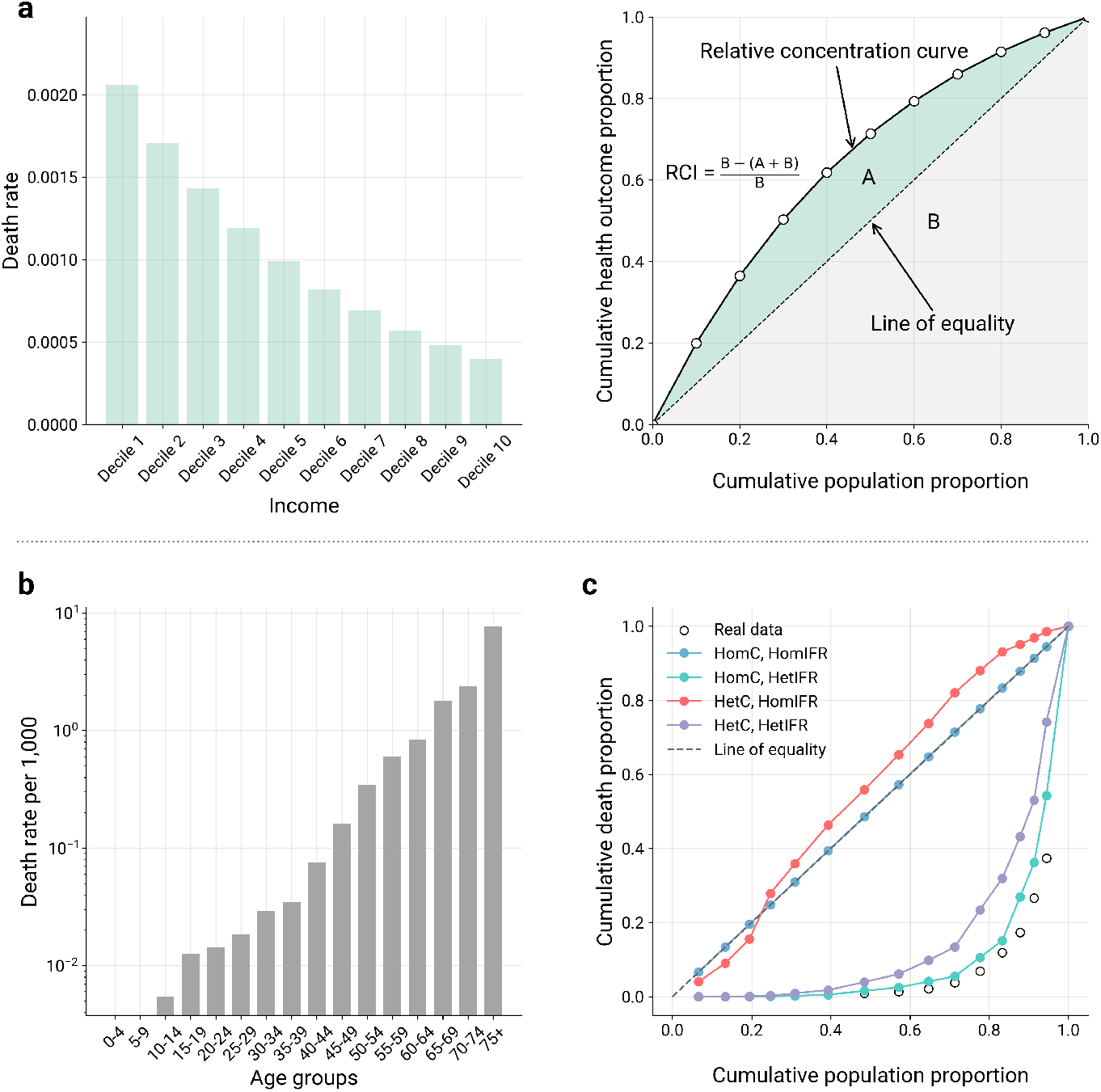
Models’ fairness and ordered social groups. **a**, The relationship between the relative concentration curve and the relative concentration index (right) obtained from a hypothetical distribution of deaths by population income deciles (left). In this example, we assume death rates to decrease as income increases. **b**, The reported COVID-19 death rates (per 1, 000) by age group in London, during the first pandemic wave between 2020*/*03*/*15 and 2020*/*07*/*05. **c**, Comparison of the relative concentration curves (RCC) of death proportion from surveillance data (open black circles) and models’ outcome, for four different epidemic models combining heterogeneous or homogeneous contact rates, and an age-dependent or flat IFR.

Given a population that can be divided into *N* ordered social groups according to a given equity-related characteristic, the RCI is computed following the formula^21^:

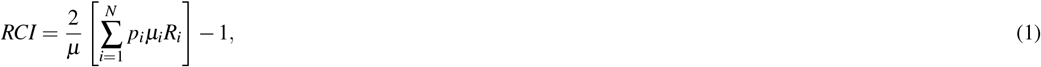

where *p*_*i*_ is the population share of group *i, µ*_*i*_ is the group average ill-health (i.e., death rate), *R*_*i*_ is the relative rank of the *i*-th group, and *µ* is the average ill-health of the whole population. The RCI ranges between −1 and 1, with extreme values corresponding to the case where the lowest (highest) ranked population group bears the full burden of the disease. Instead, an RCI = 0 indicates absolute equality between the groups. These extreme cases correspond, in Fig. 1a, to the area *A* taking values equal to *B*, − *B*, and 0, respectively.

Given the outcomes of an epidemic model and an observed distribution of the health burden related to an equity-related characteristic *s*, we propose to quantify the fairness of such a model as:

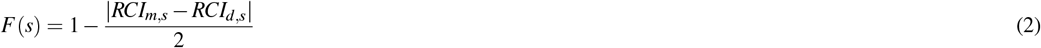

where *RCI*_*m,s*_ is the RCI of the models’ outcomes, and *RCI*_*d,s*_ is the RCI of real data when the population groups are ordered by *s. F*(*s*) ranges between 0 and 1, with 1 indicating that the model perfectly reproduces the distribution of the health outcomes observed when ordering the population groups by *s*.

We show a practical application of our definition of fairness by computing the values of *F*(*s*) for different epidemic models that describe the spread of SARS-CoV-2 in London, UK, during the first pandemic wave, between March and June 2020. In this case, we evaluate the fairness of each model in reproducing the observed inequality in the distribution of COVID-19 deaths by age. Indeed, as extensively documented, deaths from COVID-19 have been disproportionately concentrated in older age groups^30,31^. In London, the reported death rate per 1, 000 follows a highly skewed distribution, ranging from approximately 0 in infants aged 0–4 years to around 8 in individuals aged 75 and above (see Fig. 1b). This unequal distribution translates into the relative concentration curve shown in Fig. 1c (open circles), which corresponds to an *RCI*_*d,age*_ = 0.85. We explore the fairness of four age-structured SLIR-like compartmental models that combine different assumptions about the mixing patterns and infection fatality rate (IFR) adopted (see the Methods section for more details). In particular, we explore two types of mixing patterns feeding the models: homogeneous mixing across age groups (denoted as *HomC*) and synthetic age-stratified contact matrices^32,33^ (denoted as *HetC*). The models also differentiate based on the IFR used. We consider two variations: a constant IFR equal to the population average and an age-varying profile obtained from Ref.^31^. The four models correspond to the possible combinations of contact patterns and IFR. All models are calibrated to the reported total weekly deaths in London (see the Methods and Supplementary Information).

Fig. 1c shows the relative concentration curves of the median COVID-19 deaths simulated by each of the four epidemic models. As expected, the relative concentration curve produced by assuming both homogeneous mixing and a flat IFR falls on the line of equality, where the death rate is constant across groups (i.e., the absolute number of deaths is purely proportional to the group size). This model does not adequately capture the higher concentration of deaths in the elderly groups, as shown by its fairness score of *F*(*age*) = 0.58. Similarly, the model considering a flat IFR and an age-stratified contact matrix has a fairness score of *F*(*age*) = 0.53. The inclusion of an age-dependent IFR significantly improves model fairness. The model considering both age-stratified contacts and an age-dependent IFR has a fairness of *F* = 0.92 (Fig. 1b purple dots), while the one combining homogeneous mixing with a heterogeneous IFR has an even higher score of *F* = 0.98 (Fig. 1b turquoise dots). Interestingly, the inclusion of age-stratified contacts slightly worsens model fairness, both when flat and heterogeneous IFR are considered. Table S6 of the Supplementary Information summarizes these findings, showing the fairness scores based on Eq. 2, *F*(age), for the four models.

### 2.3 Fairness in unordered social groups

Next, we examine the concept of model fairness applied to the case of health inequalities associated with unordered social groups. This is the case of sociodemographic variables that do not imply any natural ordering in population subgroups, such as race and ethnicity, gender, or religious affiliation. In such a case, health inequality is generally measured through information-theoretical metrics or entropy-based indices^21^. A popular choice among Economists and Social Epidemiologists is the Theil index^34^. The Theil index measures the distribution of health among *N* population subgroups divided according to a given feature *s*, as:

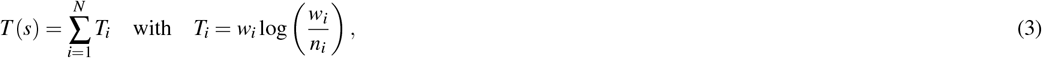

where *w*_*i*_ is the share of ill-health in group *i*, and *n*_*i*_ is the share of the population in group *i*. Higher values of the Theil index correspond to a higher degree of inequality among the groups. We propose the following definition of the fairness score for unordered social groups, based on the Theil index:

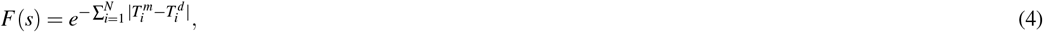

where 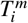 is the component of the Theil index of group *i* computed on the model’s outcome, while 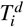 is the component of the Theil index of group *i* computed in the data. It is important to note that, in our definition of *F*(*s*), differences between the model and data are computed for each component of the corresponding Theil index (i.e., 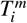 and 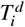) and not between the corresponding total indices. We use the exponential function to normalize *F*(*s*) in the range (0,1], with 1 indicating that a model perfectly reproduces the distribution of health among the *N* groups reported in the data.

As an illustrative example, we model the COVID-19 Pandemic in New York City during the first wave using the approach proposed by Ma et al.^35^. Based on the code shared by the authors, we develop a compartmental SLIR model that describes the spread of SARS-CoV-2 by partitioning the New York City population into five racial and ethnic groups: non-Hispanic Whites, Hispanics or Latinos, Black/African Americans, Asians, and Others. Following the approach outlined in Ref.^35^, we compare a range of model structures by integrating social contact matrices stratified by racial and ethnic groups, hypothesizing different mixing behaviors and group susceptibilities. We investigated a *variable susceptibility model*, a *proportionate mixing model*, and an *assortative mixing model*. In the *variable susceptibility model*, individuals in each group have different susceptibilities to the virus, while contact rates remain the same among the groups. In the *proportionate mixing model* and *assortative mixing model*, susceptibility is the same in all groups, while contact rates vary. Proportionate mixing assumes that the contact intensity between each pair of groups is proportional to the product of the total contact rates (i.e., total number of contacts per time period per individual) of those two groups. Assortative mixing extends proportionate mixing by partitioning a fraction *f* of contacts to be exclusively within the group while distributing the rest of the contacts according to proportionate mixing. For comparison, we define a *baseline model* with the same susceptibility and homogeneous mixing among different racial and ethnic groups. Moreover, we account for two different assumptions regarding the IFR. Heterogeneous IFRs are calculated using the age distribution within each group, providing age-adjusted IFR estimates for each group. Alternatively, we assume homogeneous IFRs for all groups. More details about the model implementation and calibration can be found in the Methods section and the Supplementary Information.

In total, we consider eight models, calibrating them to weekly COVID-19 deaths in New York City during the first wave, from March 15^*th*^ to July 5^*th*^, 2020. We then assess their fairness concerning the racial/ethnic variable using the definition given in Eq. 4. Overall, the eight models exhibit a good fit for the total reported weekly deaths, with comparable performance shown in Table S4 of the Supplementary Information. However, their ability to capture the observed disparities in death rates by race or ethnicity varies significantly.

Fig. 2 shows a comparison against the baseline of the two best models in terms of fairness (we refer the reader to Figure S7 in the Supplementary Information for the complete comparison across all models). Each panel shows the four components of the Theil index, one for each racial or ethnic group, computed from the reported COVID-19 deaths and the model outcomes. As expected, the baseline model generates an equally distributed number of deaths among racial and ethnic groups, thus failing to capture the observed inequalities. Its fairness score is *F*(*race*) = 0.714. By introducing variable susceptibility and maintaining a homogeneous IFR for all groups, the fairness of the model increases to *F*(*race*) = 0.901. A model with a homogeneous IFR and assortative mixing achieves the highest fairness *F*(*race*) = 0.906. Looking at the results for all models reported in Table S7 of the Supplementary Information, it emerges that models featuring heterogeneous IFR consistently display a lower fairness score than their counterparts with homogeneous IFR. For instance, a baseline model with equally distributed contact rates and heterogeneous IFR only achieves *F*(*race*) = 0.621 (Table S7). This suggests that the age-adjusted IFRs may not accurately reflect real-world disparities in COVID-19 death risk across racial and ethnic groups.

**Figure 2.**
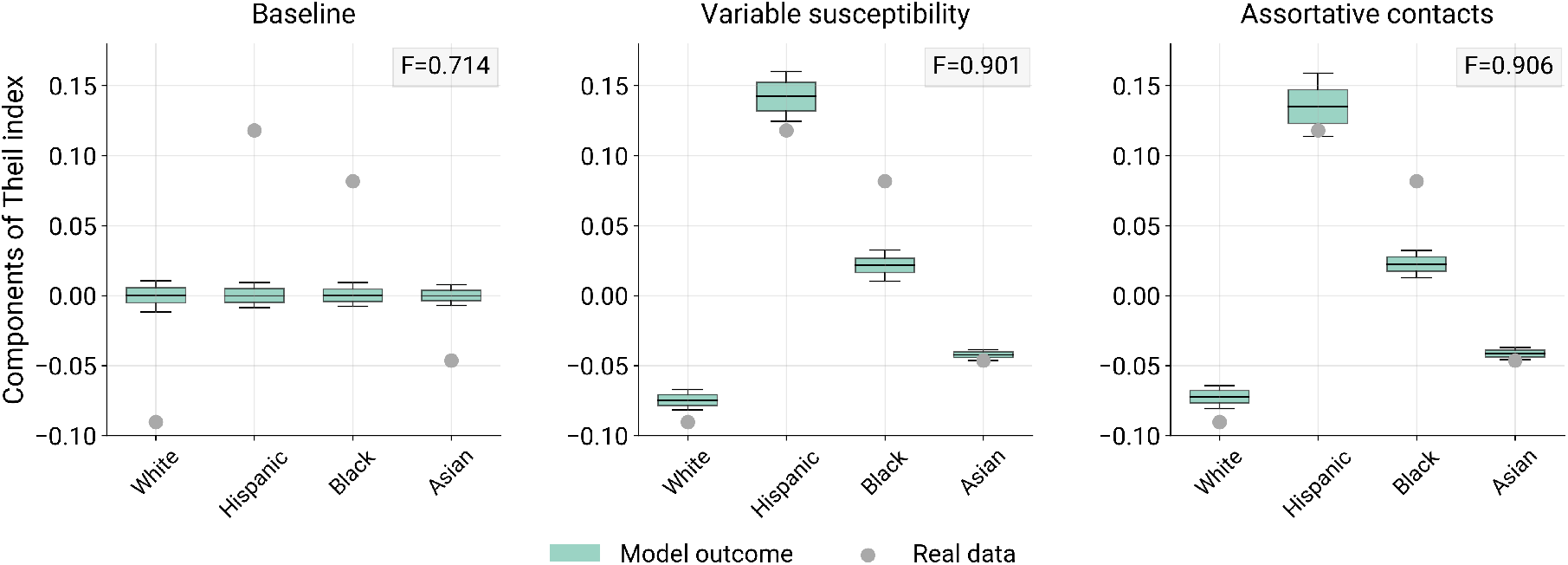
Models’ fairness for racial and ethnic groups in New York City (unordered social groups). Components of the Theil index computed from Eq. 3, according to the distribution of COVID-19 deaths by race and ethnicity in New York during the first Pandemic wave. In each panel, the Theil index components of real data are shown as gray circles. Boxplots represent the 95% reference range computed from 1, 000 simulations of each model: baseline model (left), variable susceptibility model (center), assortative contacts model (right). The corresponding fairness score of each model (i.e., Eq. 4) is shown in the top right corner.

### 2.4 Fairness in spatially structured models

As a third case, we consider spatial epidemic models. When modeling the spatial spread of an epidemic, space is typically structured into a collection of units that naturally define subgroups of the population. Such subgroups often correspond to administrative regions, and they can either induce a population ordering or not. In this case, we show how both definitions of fairness proposed above (i.e., Eq. 2 and Eq. 4) can be applied to real-world epidemic scenarios involving spatial structure.

As a first example, we examine the spread of COVID-19 in the city of Santiago, Chile, during the first pandemic wave. We consider a previously published spatially structured epidemic model that integrates human mobility patterns derived from mobile phone data^36^. The model was shown to accurately capture the unequal infection patterns observed in Santiago in 2020. Indeed, changes in mobility patterns were strongly correlated with the Human Development Index (HDI) of Santiago’s *comunas* (i.e., neighborhoods) leading to a higher burden of infection in the most deprived areas of the city (see Fig. 3a and b). To assess the role of human mobility in determining a model’s fairness, we compare the outcomes of a model that integrates human mobility data with those of a model that disregards mobility (see the Methods and Supplementary Information for more details). We use the RCC as shown in Fig. 3c, where the population of Santiago’s neighborhoods is ordered by HDI. The plot compares the cumulative proportion of COVID-19 cases reported in each community with the cumulative proportion of their population, ranked by HDI. While both models provide a good description of spatial inequalities, the RCCs show that the results of the SLIR model, which includes mobility, are closer to real data than those of the model that does not consider human movements. According to Eq. 2, the model with mobility is characterized by a fairness score of *F*(HDI) = 0.97, while excluding mobility data decreases the score to 0.93. Alternatively, we can disregard the population ordering induced by the HDI and consider each comuna of Santiago as a population subgroup. Such a grouping is unordered, and we can compute the fairness score based on the Theil index (i.e., Eq. 4) of the two models. Fig. 3d shows the components of the Theil index, one for each neighborhood, computed from the COVID-19 deaths and the average models’ outcomes. Consistent with previous results, integrating mobile phone-derived mobility data increases the model’s fairness, as evidenced by values of the Theil index components that are closer to the observed data. In this case, by including human mobility data, the overall model’s fairness increases from 0.63 to 0.79.

**Figure 3.**
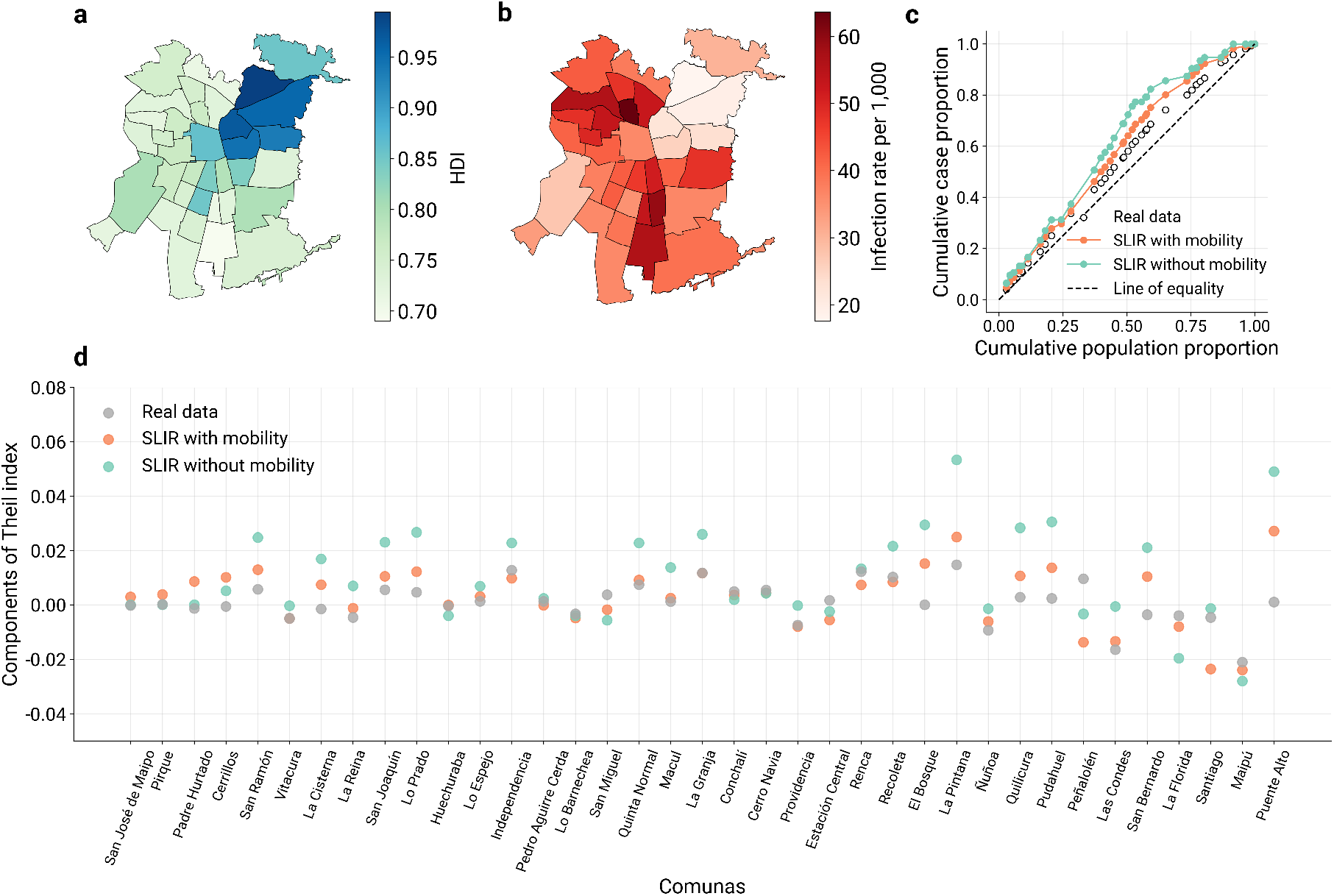
Fairness and the spatial spread of COVID-19 in Santiago, Chile (spatially structured models). **a**, Choropleth map of the urban area of Santiago, where neighborhoods (i.e., *comunas*) are color coded according to their Human Development Index (HDI). **b**, COVID-19 infection rates reported in each comuna of Santiago, during the first pandemic wave in 2020. **c**, Comparison of the relative concentration curves of COVID-19 cumulative incidence reported by the surveillance (empty circles) and generated by two models: a spatial SLIR with mobility, and the same model without mobility flows. **d**,

In the Supplementary Information, as a second example, we examine the 2015-2016 Zika virus (ZIKV) outbreak in Colombia. Again, we investigate the role of human mobility in capturing the heterogeneous burden of infections in space and compare the fairness of two modeling approaches that integrate different mobility networks. To this end, we consider the framework proposed by Perrotta et al.^37^, who modeled the spread of ZIKV in Colombia using a spatially structured model that incorporates human movements, either through mobile phone inferred mobility or through synthetic networks. Similarly to the results for Santiago, we observe that a model including mobile phone derived mobility data better captures the unequal distribution of disease across the departments of Colombia (see the Supplementary Information for more details).

We would like to stress that, although in the above examples we could consider the spatial partition of the population both as ordered and unordered, this is not always the case, and the fairness scores are not mutually exchangeable. Indeed, the RCI fairness (i.e.,Eq. 2) can only be used if an equity-related variable allows for the ordering of subpopulations on a low-high gradient. In some cases, this may not be relevant for the disease under study or may be unfeasible due to the lack of data, thus leaving the Theil index score (i.e.,Eq. 4) as the only option to evaluate a model’s fairness.

### 2.5 Fairness and equity in vaccines allocation

So far, we have discussed the concept of fairness in terms of a model’s ability to capture real-world disparities in health outcomes. However, fairness must relate not only to a model’s realism but also to the potential for improving the equity of the decisions that policymakers will make based on the model’s output. Decision making in the health domain faces significant trade-offs between algorithmic fairness constraints and the fairness of decisions^26^. Here, we show how our definition of a model’s fairness can guide the adoption of more equitable public health interventions.

To this end, we consider the COVID-19 epidemic in New York City and the model assuming assortative contacts and a homogeneous IFR, which shows the highest fairness, as described in Section 2.3. We simulate a stylized intervention scenario in which public health officials must allocate a given number of protective resources to mitigate a second wave of infections. We compare the effectiveness of different allocation strategies in terms of the achieved relative reduction in deaths compared to an unmitigated scenario. For the sake of simplicity, we assume that preventive measures consist of a sterilizing vaccine; therefore, they effectively remove individuals from the pool of susceptibles. Specifically, we consider a scenario in which policymakers must allocate *V*_*T*_ (which we assume covers 20% of the population) vaccine doses to the population, distributing them among the five racial and ethnic groups at the beginning of the second wave of the outbreak, which is triggered by the relaxation of NPIs (see Fig. 4) in July 2020. The model is calibrated and assessed in terms of fairness based on the reported COVID-19 deaths from the first wave. As for the second wave, we consider a purely hypothetical and simplified scenario obtained by simulating the relaxation of the NPIs and the emergence of a new variant with the calibrated model (see the Supplementary Information for more details).

**Figure 4.**
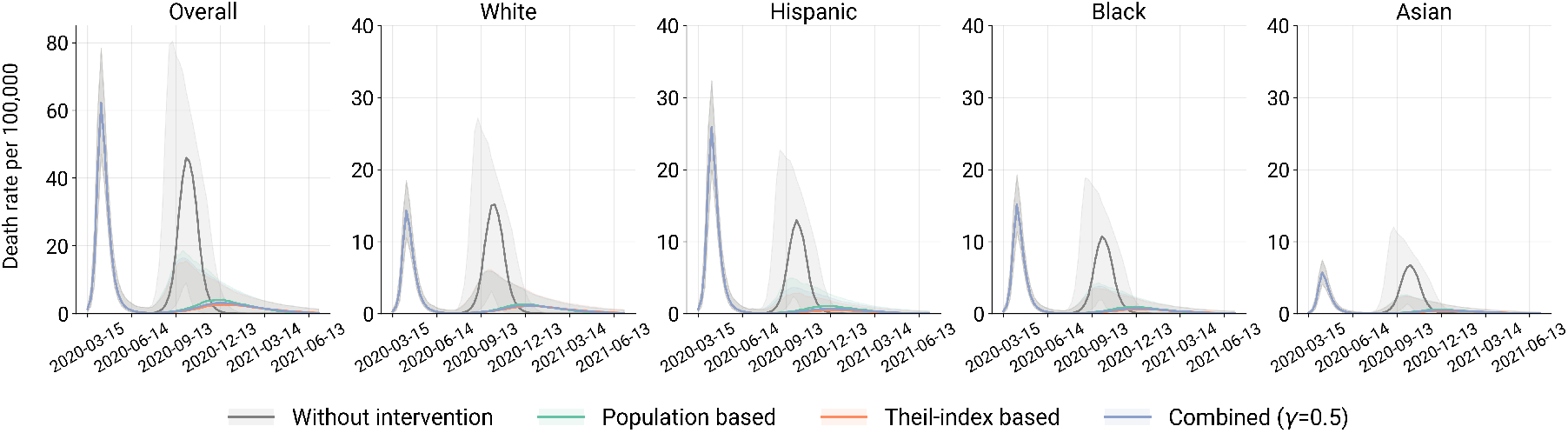
Comparison of vaccine allocation strategies on weekly deaths by race/ethnicity in NYC, given the number of vaccines *V*_*T*_ to cover 20% of the population in total. The plots show the complete deaths’ trajectories during both waves. In the first wave, there is no vaccination, thus the trajectories are overlapped. The vaccination is implemented after the first wave on 2020*/*07*/*06 in all scenarios. In the plot, we show the median of weekly deaths with 95% confidence interval of 1, 000s samples.

Vaccine allocation can be done by assigning vaccines to each group proportionally to the relative size of the group population *n*_*i*_. This is our baseline scenario that leads to an overall 68.2%[53.8% −94.2%] relative reduction in the number of deaths (see Table 1). Alternatively, we consider allocating the *V*_*T*_ doses using a formula that weights the population share of each group by the Theil index component introduced in Eq. 4. In this way, we directly introduce the fairness metric into the allocation, effectively linking our measure of the model’s fairness to the decision-making process. Specifically, vaccines are distributed to each group according to their corresponding values of 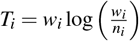, where *w*_*i*_ is the fraction of deaths reported by group *i* during the first wave. We then introduce *T*_*i*_ into the vaccine allocation, controlling the relative importance of the Theil index through a parameter *γ* ranging between 0 and 1 (see the Methods for more details). For *γ* = 1, we implement a strategy that is fully based on the Theil index components, while smaller values of *γ* take into account the group population size.

**Table 1.** Comparison of relative reduction in the number of deaths, by vaccine allocation strategy and racial/ethnic groups. The most effective allocation strategy, overall and for each population subgroup, is highlighted in bold. Each entry reports the median reduction from 1, 000 samples and the 95% CI.

| Strategy | Relative reduction in deaths (%) |  |  |  |  |
| --- | --- | --- | --- | --- | --- |
|  | Overall | White | Hispanic | Black | Asian |
| Population based | 68.2 [53.8, 94.2] | 68.9 [54.3, 94.4] | 69.2 [54.7, 94.8] | 67.7 [52.9, 94.3] | 69.7 [55.0, 94.4] |
| Theil-index based | **73.9 [56.6, 97.9]** | 69.2 [49.7, 97.6] | **82.5 [68.7, 98.7]** | **73.2 [55.6, 97.9]** | **72.5 [53.9, 97.8]** |
| Combined ($\gamma = 0.5$) | 71.6 [55.3, 96.9] | **69.4 [52.2, 96.6]** | 76.8 [62.2, 97.8] | 70.8 [54.3, 97.0] | 71.3 [54.8, 97.0] |

Table 1 compares the relative reduction in deaths achieved by different vaccine allocation strategies based on the components of the Theil index for each racial/ethnic group. In general, allocation strategies based on the Theil index achieve a larger overall reduction in deaths and, at the same time, improve outcomes for all racial and ethnic groups compared to the baseline. By integrating the Theil index components into the allocation strategy, the Theil-index based strategy (*γ* = 1) and a combined strategy (*γ* = 0.5) achieve an overall median reduction of 73.9% and 71.6% in the number of deaths, respectively, with an increased reduction in deaths across all groups relative to the baseline. The Theil-index based strategy consistently achieves the largest reduction in overall deaths when varying the value of *V*_*T*_ between 10% and 30% (see Tables S9 and S10 of the Supplementary Information).

## 3 Discussion

We proposed a set of metrics to quantify and compare the fairness of models of infectious disease dynamics. Our goal was to bring established principles and measures from Social Epidemiology^38^ into Computational Epidemiology, thereby bridging the two research areas that have remained largely disconnected. By contrasting different modeling approaches and disease settings, we showed that our fairness metrics can measure a model’s ability to capture the observed inequalities in the distribution of illness between sociodemographic groups. Our results highlighted trade-offs between model complexity and fairness. We demonstrated that increasing model complexity through additional data layers or compartments, such as the presence of heterogeneous contact rates between groups, does not always enhance its fairness. Similarly, models that achieved the best overall fit to disease data did not translate into the fairest outcomes when examining inequalities between sociodemographic groups. Therefore, our findings point to the idea that maximizing the fairness of an epidemic model cannot be simply achieved through total error minimization or by increasing models’ complexity. Our fairness metrics offer an alternative to traditional accuracy measures by aligning more closely with the underlying concept of health inequality and its measures. Indeed, as widely accepted in the field of Machine Learning and more broadly in Artificial Intelligence, algorithmic fairness goes beyond mere accuracy, placing greater emphasis on ensuring equitable outcomes^39^. To address this important point, we demonstrated that the definition of fairness in our framework provides a useful score to increase the equity of a hypothetical intervention, such as vaccine allocation during an outbreak. Indeed, our results show that when introduced as a guide for a vaccine allocation strategy, our measure of fairness reduces the overall disease burden while increasing equity in disease outcomes between groups. This effectively addresses the trade-off between equity and mortality^40^.

Algorithmic fairness in the health domain, especially when related to clinical practice, represents a highly debated topic that has been extensively investigated^25,41^. In public and population health, algorithmic fairness has gained attention more recently, and its definition remains elusive as it intersects with complex social determinants, systemic inequities, and the limitations of available data^17^. For computational epidemic modeling, the debate is still in its infancy. Previous studies have examined the fairness of COVID-19 forecasting models in the USA^18–20^, but they have not accounted for global measures of health disparities across subgroups, nor for ways to evaluate the fairness of policy decisions. Our study provides epidemic modelers with a principled and more general framework that can be easily adapted to different contexts and modeling approaches, as demonstrated by the examples we considered.

Our work is subject to limitations. First, we did not address the important issue of correctly identifying, from available data, the equity-related variables that must be included in an epidemic model and how a model can account for the multiple intervening mechanisms of disease exposure associated with one or more sociodemographic traits^42^. Indeed, the models we considered did not attempt to mechanistically link social causes with biological outcomes. This may explain why models showed low fairness scores in some settings, such as the ZIKV epidemic in Colombia. We expect that future modeling advances will improve the state-of-the-art through novel approaches that incorporate social drivers of inequity at their core, thus increasing models’ fairness. Second, as already noted, we considered a simplified vaccine allocation scenario without aiming for realism. For instance, we did not address the ethical implications arising from a limited vaccine supply or the existence of adverse impacts from the vaccine^43^. To effectively evaluate the relevance of our fairness metrics for decision making, a more extensive analysis is needed across different epidemic and intervention scenarios. This will be the subject of future work.

Computational epidemiologists and mathematical modelers have recently begun to address the socioeconomic inequalities that characterize the spread of infectious diseases^10,13^. The COVID-19 pandemic has demonstrated that inequalities in epidemic outcomes are strongly associated with socioeconomic disparities, systemic racism, and (lack of) social justice^44–46^. Such inequalities were reflected by significant disparities in social contacts^47,48^, the affordability of non-pharmaceutical interventions^49–52^, access to healthcare^5^, and job security^53^. To address these pressing issues, emerging literature has put forward novel approaches to integrate social mechanisms of exposure into traditional epidemic models, such as through generalized contact matrices^14,54^ or the use of structural causal influence^55^. Our work adds to this current thread of research by providing modelers and public health officials with a new framework that will help evaluate epidemic modeling outcomes through the lens of equity, thus reducing socioeconomic biases in outbreak management and response.

## 4 Methods

### Epidemic models

In all cases, we considered computational epidemic models that are fully stochastic and simulated using chain binomial processes. In the following, we provide the details of each model structure, parameters, and underlying assumptions.

#### Simulating the spread of SARS-CoV-2 in London

We use a standard age-structured SLIRD (Susceptible, Latent, Infectious, Recovered, Dead) compartmental model incorporating the impact of non-pharmaceutical interventions (NPIs). Individuals are divided into compartments according to their health status. The transitions between compartments are as follows. Susceptible (*S*) individuals transition to the latent compartment after interacting with infectious individuals. Individuals in the latent (*L*) phase are infected but not infectious until they move to the infectious (*I*) phase after the latent period. Then, infectious individuals either die (*D*) with an infection fatality rate (*IFR*) or recover (*R*) from the disease at a rate of 1 − *IFR*. To account for a delay in reporting deaths, we set a parameter of Δ days of delay, and the individuals in the *D* compartment are moved to *D*^*o*^ after Δ days. In calibration, the number of individuals in *D*^*o*^ is calibrated against the reported deaths.

The population is stratified into 16 age groups from 0 to 75+, with 5-year brackets except for the 75+, with age-specific contact patterns and IFRs. We consider two types of mixing among age groups: 1) homogeneous and 2) heterogeneous mixing. In the models with homogeneous mixing, we assume that the number of contacts between any two age groups is proportional to the product of the populations in those two groups. Based on this assumption, the homogeneous matrix is constructed by redistributing the total contacts derived from Ref.^32^ to each pair of age groups (see Section 1.2 in the Supplementary Information for more details). In the models with heterogeneous mixing, contacts between age groups are described by contact matrices from Prem et al.^32^. To include the impact of NPIs, we use mobility data from the Google COVID-19 Community Mobility Report^56^ and the Oxford Coronavirus Government Response Tracker (OxCGRT)^57^ to estimate the reduction in overall contacts and adjust the contact matrices (see Section 1.1 in the Supplementary Information). We also consider two approaches to model deaths: 1) homogeneous and 2) heterogeneous IFRs. In the models with homogeneous IFR, infection fatality rates are the same across the age groups, and their values are computed as the population-weighted average of the age-specific values. In the models with heterogeneous IFR, we use the age-specific IFRs^31^.

#### Simulating the spread of SARS-CoV-2 in New York City

Here, we adopt and extend the modeling framework proposed by Ma et al^35^. While it shares many ingredients of the model we used in the case of London, such as the integration of the effects of NPIs, there are also some important differences. Indeed, instead of considering sub-groups by age, the population is divided into five racial and ethnic groups: non-Hispanic Whites, Hispanics or Latinos, Black/African Americans, Asians, and Others. Ma et al proposed three models: the *variable susceptibility model*, the *proportionate mixing model*, and the *assortative mixing model*^35^. We adopt and extend these, building eight models by varying the assumptions regarding susceptibility, contact patterns, and IFR. In the *variable susceptibility model*, individuals from each racial/ethnic group have different susceptibilities to the virus, while the contact rates are identical across all groups. In the *proportionate mixing model*, individuals have the same susceptibility, while the contact rate between a pair of groups *i* and *j* is proportional to the product of the total contact rates of group *i* and *j*. In the *assortative mixing model*, the contact pattern is an extension of proportionate mixing, partitioning a fraction *f* of contacts to be exclusively within the group while distributing the rest of the contacts according to proportionate mixing. In addition to the above three models, we propose a baseline seeded with identical susceptibility across racial and ethnic groups, assuming a homogeneous mixing assumption. Then, we double the above four models by considering two alternative IFRs (homogeneous or heterogeneous), which results in a total of eight models (see Section 1.3 in the Supplementary Information for more details). We adopt the calibrated susceptibility estimates reported in Ma et al.^35^ for the models with variable susceptibility and set susceptibility to 1 for the models assuming equal susceptibility. The contact matrices are also estimated from Ref.^35^. For each model, the corresponding formulations of the contact matrices and IFR are provided in the Supplementary Information.

#### Spatially structured metapopulation models

To simulate the spread of COVID-19 in Santiago de Chile and the spread of ZIKV in Colombia, we use two spatially structured metapopulation models that integrate human movement patterns and disease dynamics^58,59^. In both models, space is divided into patches corresponding to administrative subdivisions (i.e., neighborhoods in Santiago and departments in Colombia), which are connected by mobility fluxes. In each patch, individuals interact according to the specific compartmental model that describes the disease under study. Both models are stochastic, and transitions among compartments are simulated through chain binomial processes.

In the case of Santiago, the results presented in this work are based on the data and simulations published in Ref.^36^. All the details regarding the model implementation, calibration, and analysis can be found therein. The model for Colombia, instead, is extensively described in Ref.^37^. The results presented in this work are based on the data and simulations published therein.

#### Models calibration

We calibrate the models for the cases of London and NYC by fitting the aggregated weekly deaths against the reported data. We use the Approximate Bayesian Computation Sequential Monte Carlo (ABC-SMC) algorithm^60^ for calibration. The ABC-SMC algorithm is an extension of the ABC rejection method that utilizes generated intermediate distributions and decreasing rejection thresholds. The algorithm consists of *T* generations, and within each generation, *M* particles (sampled parameters) are maintained. In the first generation, parameters *θ*_*i*_ (*i*=1, 2, …, *M*) are sampled from the prior distribution *π*(*θ*), and *θ*_*i*_ are accepted if the distance 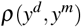 between the observed data *y*^*d*^ and the simulated results *y*^*m*^ ∼ *f* (*θ*_*i*_) is smaller than an initial threshold *ε*_1_. In the following generations *t* (*t >* 1), particles are sampled from the previously accepted generation 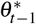 and perturbed by a kernel *K*(*θ*^*^). They are accepted if the same distance *ρ*(*y*^*d*^, *y*^*m*^) falls below *ε*_*t*_, where *ε*_*t*_ is the threshold of generation *t* and *ε*_*t*_ *< ε*_*t*−1_. We adopted a Python implementation of ABC-SMC from the library *pyabc*^61^. In our simulations, we set *T* = 10, *M* = 1000, and we take the last accepted generation as the posterior distribution of the parameters in our model. The settings for free parameters, along with their prior distributions and posterior distributions, are presented in the Supplementary Information.

#### Fairness and vaccine allocation strategies

We consider a counterfactual intervention scenario based on the model of COVID-19 spread in New York City. Specifically, we assume that public health officials allocate a given number of vaccines (*V*_*T*_) to the population at the beginning of the second wave of the outbreak. At the end of the first wave, which is calibrated to the reported COVID-19 deaths, we simulate a second wave by assuming the emergence of a new variant with a 50% higher transmission rate. We also assume that the relaxation of NPIs leads to higher contact rates with respect to the last day of the first wave (see section 4.1 in the Supplementary Information for more details). For simplicity, we assume that only susceptible individuals are vaccinated and that the vaccines provide sterilizing immunity. In other words, we remove a number of *V*_*T*_ individuals from the pool of the susceptible, and they cannot be infected. In this setting, we investigate three vaccine allocation strategies:

- *Population-based strategy*. Vaccines are distributed to each group proportionally to the population size of that group. The number of vaccines allocated in group *i* (denoted by *V*_*i*_) is 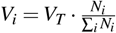
- *Theil index-based strategy*. We consider both the population size and the Theil index component of each group for vaccine distribution. The number of vaccines distributed to group *i* is

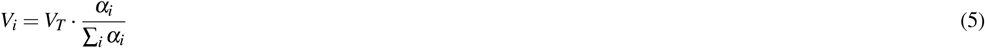

where we define *α*_*i*_ as

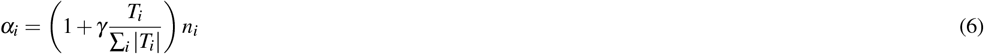

where *T*_*i*_ is the component of the Theil index of group *i* (Eq. 3), and *γ* is a parameter controlling the importance of Theil index components, ranging in [0, 1]. If *γ* = 1, the importance of the Theil index is maximized, and we define it as the full Theil based strategy. If *γ* = 0, the strategy is reduced to a population-based strategy. Intermediate values of *γ* define allocation strategies that mix the population distribution and the Theil index components.

## Acknowledgements

We thank Daniela Perrotta for sharing simulation results from her previously published work, which were helpful in developing the results presented in this paper. We gratefully thank Alessandro Vespignani for providing useful comments on an early version of the manuscript. M.T. thanks the mathematical research institute MATRIX in Australia, where part of this research was initially conceived.

## Data availability statement

Data and code to reproduce the study are available on GitHub: https://github.com/Jadecoool/Fairness-in-computation

## Author contributions statement

M.T. conceived the project; N.G., N.P., and M.T. designed the study and supervised the research; Y.L. investigated and visualized the results; Y.L., N.G., N.P., and M.T. contributed to the methodology; M.T. contributed to the writing — original draft; Y.L., N.G., N.P., and M.T. contributed to the writing, review, and editing.

