## Supplementary Information for "Fairness in infectious disease modeling"

November 11, 2025

### 1 Model Descriptions

Here, we provide the details of the epidemic models used in the case of ordered social groups and unordered social groups. All models are based on the classic Susceptible-Latent-Infected-Recovered (SLIR) compartmental modeling framework. Furthermore, they incorporate the impact of the implementation of non-pharmaceutical interventions (NPIs).

We consider 16 age groups from 0 to 75 with a 5 year bracket, except for the last group of 75+. Each age group follows the epidemic transmission dynamics described below. Susceptible individuals ($S$) transition to the latent stage ($L$ compartment) at a rate $\lambda$. This is the force of infection which, in general, is a function of the density of infectious individuals, contact matrix, the transmissibility $\beta$ of the virus, and NPIs. Individuals in the latent stage are infected but not yet infectious. They stay in $L$ for an average of $\epsilon^{-1}$ $days^{-1}$. Afterwards, they become infectious and enter the $I$ compartment. After an average of $\mu^{-1}$ $days^{-1}$, they either recover from the disease (transitioning to $R$) with a probability $1 - IFR$ or die from the disease with a probability $IFR$, where $IFR$ is the infection-fatality rate. We also consider a delay of $\Delta$ days in death reporting. After $\Delta$ days, we remove new deaths from compartment $D$ moving them to $D_o$.

#### 1.1 Models for ordered social groups in London

We investigate four models that differ in their assumptions regarding contact matrices and infection fatality rates. We consider two types of contact matrices (i.e., homogeneous/heterogeneous) and two types of IFRs (i.e., homogeneous/heterogeneous). Accordingly, we consider all possible combinations of contact matrices and IFRs, which result in four models featuring:

1. homogeneous contacts and homogeneous IFR (HomC-HomIFR)
2. homogeneous contacts and heterogeneous IFR (HomC-HetIFR)
3. heterogeneous contacts and homogeneous IFR (HetC-HomIFR)
4. heterogeneous contacts and heterogeneous IFR (HetC-HetIFR)

We use the contact matrices from Ref. [1]. Specifically, we consider contact matrices at four locations: home $C_{ij}^{home}$, $C_{ij}^{work}$, $C_{ij}^{school}$, and other places $C_{ij}^{other}$, where the $i$th and $j$th entries indicate the contact rate of group $i$ with group $j$. We take the sum of the four matrices as the heterogeneous contact matrix in our model, denoted by $C_{ij}^{Het}$. It is computed as

$$C_{ij}^{Het} = C_{ij}^{home} + C_{ij}^{work} + C_{ij}^{school} + C_{ij}^{others} \tag{1}$$

Homogeneous contacts assumes that the number of contacts between two groups is proportionate to the product of the population size of those two groups. We compute the homogeneous contact matrix as follows. First we obtain the total number of contacts $\mathcal{R}$ through the heterogeneous contact matrix by $\mathcal{R} = \sum_{i,j} C_{ij}^{Het} \cdot N_i$, where $N_i$ is the population of group $i$. According to the assumption of homogeneous mixing, we assign the number of contacts to a pair of group $i$ and $j$ proportionately to the fraction $\frac{N_i N_j}{N^2}$, where we defined $N = \sum_i N_i$. Hence, the homogeneous contact matrix $C_{ij}^{Hom}$ is computed as the number of contacts of group $i$ with group $j$ divided by the population size of group $i$:

$$C_{ij}^{Hom} = \frac{1}{N_i}\mathcal{R} \cdot \frac{N_i N_j}{N^2} = \frac{\mathcal{R}}{N^2} N_j \tag{2}$$

The heterogeneous IFRs are set according to Ref. [2]. While the homogeneous IFRs are identical across age groups, computed as the average value of the heterogeneous IFRs weighted by the population size of each group:

$$IFR_{Hom} = \frac{N_i}{\sum_i N_i} IFR_i \tag{3}$$

where $IFR_i$ is the heterogeneous IFR of group $i$.

We simulate the models by using stochastic chain binomial processes. For age group $i$, the compartmental model is defined by the following set of stochastic equations:

$$S_i(t + \delta t) = S_i(t) - Bin(S_i(t), \lambda_i) \tag{4}$$

$$L_i(t+\delta t) = L_i(t) + Bin_1(S_i(t), \lambda_i) - Bin(L_i(t), \epsilon) \tag{5}$$

$$I_i(t+\delta t) = I_i(t) + Bin(L_i(t), \epsilon) - Bin(I_i(t), \mu) \tag{6}$$

$$R_i(t+\delta t) = R_i(t) + Bin(I_i(t), \mu)(1 - IFR_i) \tag{7}$$

$$D_i(t+\delta t) = D_i(t) + Bin(I_i(t), \mu) IFR_i \tag{8}$$

$$D_{oi}(t+\delta t) = D_{oi}(t+\delta t - \Delta) \tag{9}$$

$$\tag{10}$$

where the force of infection is

$$\lambda_i = \beta \sum_j \frac{C'_{ij} I_j}{N_j} \tag{11}$$

and $C'_{ij}$ is the adjusted contact matrix by NPIs (see section 1.3) based on either $C_{ij}^{Hom}$ or $C_{ij}^{Het}$. $I_j$ is the number of infected individuals in group $j$. $\beta$ is the transmission rate of the virus.

#### 1.2 Models for unordered social groups in NYC

We investigate eight models with different assumptions regarding susceptibility, contact matrices, and IFRs across racial/ethnic groups. We first investigate three models proposed in Ref. [3]: the ***variable susceptibility model***, ***proportionate mixing model***, and the ***assortative mixing model***. While we introduce the three models briefly, we refer the reader to Ref. [3] for more information. In the ***variable susceptibility model***, individuals of each group have different susceptibility **q** to the virus, while contact rates are the same among groups. In the ***proportionate mixing model*** and ***assortative mixing model***, susceptibility is identical among groups, while contact rates are different. Proportionate mixing assumes that the contact intensity for each pair of two groups is proportional to the product of the total contact rates of those two. Assortative mixing extends the proportionate mixing by partitioning a fraction $f$ of contacts to be exclusively within the group while distributing the rest of the contacts according to proportionate mixing.

We construct the contact matrices following Ref. [3]. The contact matrix of the ***variable susceptibility model*** is homogeneous, as described in Eq. 2. The contact matrix of the ***proportionate mixing model*** is computed as:

$$C_{ij}^{prop} = \frac{a_i a_j N_j}{\sum_k a_k N_k} \tag{12}$$

where $N_j (N_j \text{ or } N_k)$ is the population size of group $i(j \text{ or } k)$. $a_i (a_j \text{ or } a_k)$ is the total contact rate of group $i(j \text{ or } k)$, i.e., the number of contacts per person in group $i(j \text{ or } k)$. The values of $a_i$ are taken from Ref [3].

The contact matrix of the ***assortative mixing model*** is computed as:

$$C_{ij}^{assort} = (1-f) \frac{a_i a_j N_j}{\sum_k a_k N_k} + f \delta_{ij} a_i \tag{13}$$

where $N_j (N_j \text{ or } N_k)$ is the population size of group $i(j \text{ or } k)$. $a_i (a_j \text{ or } a_k)$ is the total contact rate of group $i(j \text{ or } k)$, i.e., the number of contacts per person in group $i(j \text{ or } k)$. $f$ is the fraction of contacts to be exclusively within-group. The values of $a_i$ and $f$ are taken from Ref. [3].

For comparison, we establish a ***baseline model*** with homogeneous assumptions for susceptibility and contacts. Specifically, we consider the same susceptibility across racial/ethnic groups and homogeneous mixing with the contact matrix calculated through Eq. 2.

In addition to different assumptions for susceptibility and contact matrices, we account for two types of IFRs (i.e., heterogeneous/homogeneous). The heterogeneous IFRs are age-adjusted for racial/ethnic groups, calculated using the age distribution within each group. The IFR of group $i$ is computed as

$$IFR_i = \sum_k \frac{N_{ik}}{N_i} IFR_k \tag{14}$$

where $IFR_k$ is taken from age-specific IFRs from Ref. [2]. $N_i$ is the population in group $i$, $N_{ik}$ is the population of age group $k$ of group $i$. The homogeneous IFR assumes the same value across racial/ethnic groups, computed as the weighted average value of the heterogeneous IFRs according to Eq. 3.

Under the two IFR configurations, we extend the ***baseline model***, ***variable susceptibility model***, ***proportionate mixing model***, and the ***assortative mixing model*** into two versions each (with homogeneous/heterogeneous IFR), which results in 8 models in total, labeled as:

1. baseline model with homogeneous IFR (Baseline-HomIFR)
2. variable susceptibility model with homogeneous IFR (Suscept-HomIFR)
3. proportionate mixing model with homogeneous IFR (Prop-HomIFR)
4. assortative mixing model with homogeneous IFR (Assort-HomIFR)
5. baseline model with heterogeneous IFR (Baseline-HetIFR)
6. variable susceptibility model with heterogeneous IFR (Suscept-HetIFR)
7. proportionate mixing model with heterogeneous IFR (Prop-HetIFR)
8. variable susceptibility model with heterogeneous IFR (Assort-HetIFR)

Analogously to what done in the case of ordered social groups, the dynamics of models for unordered social groups are also described by stochastic chain binomial processes, following Eqs. 4-1.1.

#### 1.3 Implementation of non-pharmaceutical interventions

In both cases of ordered/unordered social groups, we model the non-pharmaceutical interventions (NPIs) by using mobility data. We assume that NPIs have a homogeneous impact on mobility levels across subgroups. To this end, we use the COVID-19 Community Mobility Report released by Google [4] to re-scale the contact matrices as follows:

$$\mathbf{C}' = \left(1 + \frac{p}{100}\right)^2 \cdot \mathbf{C} \tag{15}$$

where $p$ is the average percentage of change in mobility at home ($p_{home}$), work ($p_{work}$), and other places ($p_{others}$) with respect to a pre-pandemic baseline measured by Google [4]. Specifically, $p_{home}$ refers to the changes in household contacts, $p_{work}$ indicates workplace contacts, and $p_{others}$ represents the contacts in retail stores, recreation sites, and transit stations. We assume a square form, as the number of contacts is proportional to the square of the number of individuals in a location [5, 6]. Negative values of $p_{home}/p_{work}/p_{others}$ indicate a decrease in mobility, while positive values indicate an increase in mobility with respect to the pre-pandemic mobility levels.

#### 1.4 Model parameterization and calibration

We use the Approximate Bayesian Computation Sequential Monte Carlo (ABC-SMC) algorithm [7] to calibrate the models. The goal is to estimate the posterior distribution for models' parameters given the input of their prior distribution. The algorithm consists of $T$ generations (i.e., iterations). The first generation is based on a rejection algorithm step where the tolerance $\xi$ is set to a high value. In the second generation, the tolerance is decreased, parameters are sampled from those accepted in the previous step and perturbed via a kernel to avoid converging on local minima of the phase space. The process is repeated for $T$ generations of $M$ particles (i.e, samples) each. Then, the set of accepted parameters in the last generation is used as the empirical posterior distribution. We adopted a python implementation of ABC-SMC from the library *pyabc* [8]. In our simulations, we set $T = 10$ and $M = 1000$. We use the Weighted Mean Average Percentage Error (wMAPE) as the metric for measuring the distance between the simulated trajectory of a model and the reported data. It is computed as:

$$\text{wMAPE} = \frac{\sum_t |y_t^d - median(y_{i,t}^m)|}{\sum_t y_t^d} \tag{16}$$

where $y_t^d$ is the reported data, $median(y_{i,t}^m)$ is the median trajectory of sample $i$. Both are 1-dimensional vectors of time $t$.

For the models of ordered social groups in London and unordered social groups in NYC, we explore the same free parameters as follows. The priors of these parameters are:

- Reproductive number $R_0$. We explore values in a uniform interval $1.5 - 6$.
- Delay in reporting deaths $\Delta$. Consistent with observations, we explore integers in a uniform interval $7 - 35$.
- Initial number of infected individuals $I_0$. We explore integers in a uniform interval $1 - 10000$.

The initialization of the models is as follows. For both cases of London and NYC, we simulate the first wave of COVID-19 from $2020-03-15$ to $2020-07-05$. At the beginning, there are $I_0$ infected individuals. We assign $I_0$ to compartment $L$ and $I$ proportionately to their periods, $\frac{\epsilon^{-1}}{\epsilon^{-1}+\mu^{-1}}$ or $\frac{\mu^{-1}}{\epsilon^{-1}+\mu^{-1}}$ respectively. Then, for each group $i$, we assign the number of individuals in $L$ or $I$ proportional to the population size of each group. Therefore, the initial number of individuals in $L$ of group $i$ is

$$L_i(t=0) = I_0 \cdot \frac{\epsilon^{-1}}{\epsilon^{-1}+\mu^{-1}} \cdot \frac{N_i}{N}, \tag{17}$$

the initial number of individuals in $I$ of group $i$ is

$$I_i(t=0) = I_0 \cdot \frac{\mu^{-1}}{\epsilon^{-1}+\mu^{-1}} \cdot \frac{N_i}{N} \tag{18}$$

where $i_0$ is the total number of infected individuals among the whole population, $N$ is the total population size, $N_i$ is the population size of group $i$.

We assume all other individuals are susceptible at the beginning of the epidemic. Thus, $S_0 = N - I_0$. In the same way, for each subgroup, we distribute $S_0$ proportionately to the population size in each group:

$$S_i(t=0) = S_0 \cdot \frac{N_i}{N} \tag{19}$$

All parameters of the models are presented in Table S1.

**Table S1.** Model parameters.

| **Notation** | **Description** | **Value** |
|---|---|---|
| $R_0$ | Reproductive number | Calibrated within $1.5 - 6$. |
| $\Delta$ | Delayed days in reporting deaths | Calibrated within $7 - 35$ |
| $I_0$ | Initial number of infections | Calibrated within $1 - 10000$ |
| $\epsilon$ | Latent period | $3.7 days$ sourced from Ref. [9,10] |
| $\mu$ | Infectious period | $2.5 days$ sourced from Ref. [9,11] |
| $\beta$ | Transmissibility | Computed from $R_0$ |

The posteriors of the free parameters in the ordered social group case and the unordered social group case are shown, respectively, in Figs. S1 and S2. We show the median along with 50% confidence interval (CI) marked by darker boxes, and 95% CI marked by lighter boxes of the selected $1,000$ samples. In the case of London, the median of the reproductive number $R_0$ falls in the range [1.70, 1.96] across the four models. In the case of NYC, the median reproductive number $R_0$ falls in the range [2.17, 2.24] across the eight models. These results align with previous findings [12–14]. For the delay in reporting deaths $\Delta$ and the initial number of infected individuals, both cases exhibit similar posteriors across all models.

### 2 Models Fit

In this section, we present the model fits to both aggregated and group-level data.

#### 2.1 Fitting of COVID-19 deaths in London

We fit the models to COVID-19 weekly deaths in London during the first wave, spanning from $2020-03-15$ to $2020-07-05$. Fig. S3 shows the aggregated deaths' trajectories. All models are able to fit the data. To quantitatively compare the models' outcomes, we compute the wMAPE of the median trajectory of the

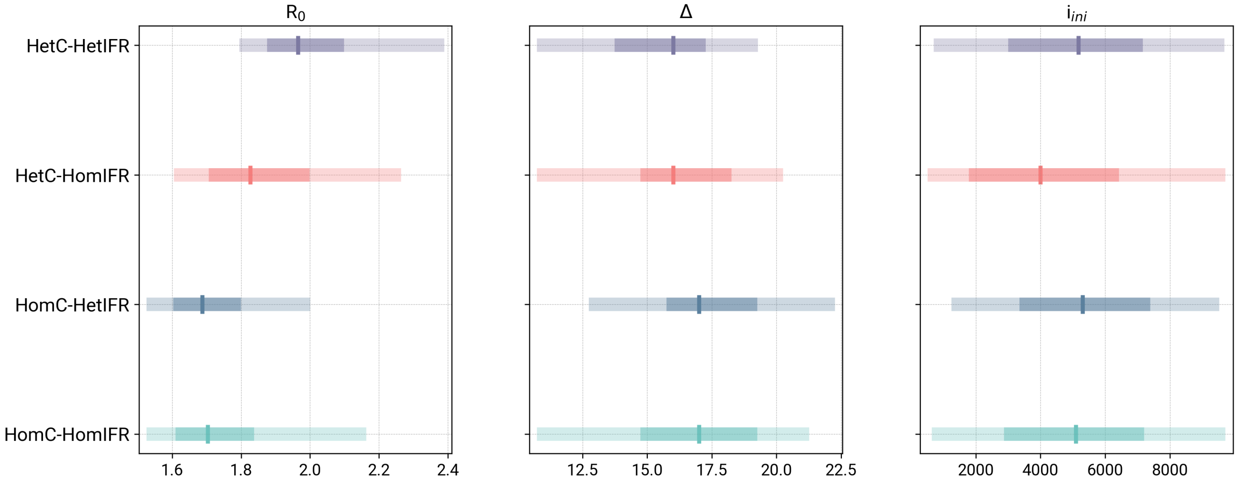

**Fig S1. Posteriors of free parameters of the models for ordered social groups in London.**

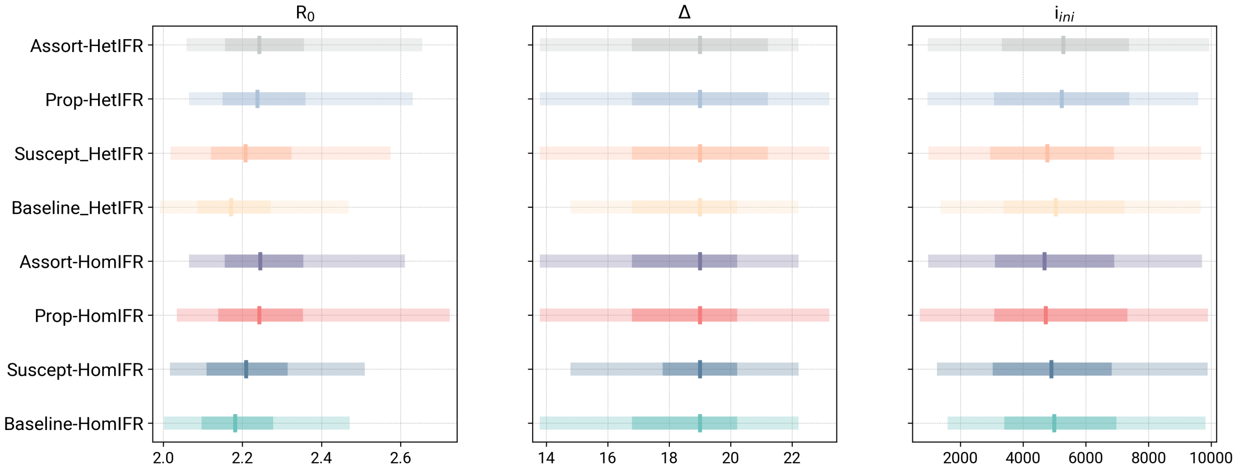

**Fig S2. Posteriors of free parameters of the models for unordered social groups in NYC.**

models. As shown in Tab. S2, the HetC-HomIFR model shows the lowest wMAPE, indicating the best fitting to the reported deaths. In contrast, the HomC-HetIFR yields the worst wMAPE.

To investigate the capability of models in fitting the inequality, we present the fit of the models to weekly deaths at the group level. As shown in Fig. S4, we can see differences in the models' performances. The two models with heterogeneous IFR (HomC-HetIFR, HetC-HetIFR) show better fits in age groups $0-4$ to $45-49$ compared to the other two models with homogeneous IFR (HomC-HomIFR, HetC-HomIFR). In terms of the 75+ age group, the HomC-HetIFR model provides the best fit to the real data, followed by the HetC-HetIFR model. In order to compare the models quantitatively, for each model we compute the wMAPE between models' outcomes and the reported data for each subgroup, and take the median value of the wMAPEs across subgroups. The results are shown in Tab. S3. The HomC-HetIFR model shows the lowest median wMAPE, indicating the best performance in fitting group-level weekly deaths. In contrast, the HetC-HomIFR model reports the worst median wMAPE. Overall, the performance of the models in fitting the aggregated death trajectory is similar; however, their performances vary in fitting the subgroup-level data.

**Table S2.** wMAPEs of simulated aggregated weekly deaths of models with respect to reported aggregated deaths in London

| HomC-HomIFR | HomC-HetIFR | HetC-HomIFR | HetC-HetIFR |
|---|---|---|---|
| 0.143 | 0.146 | **0.121** | 0.141 |

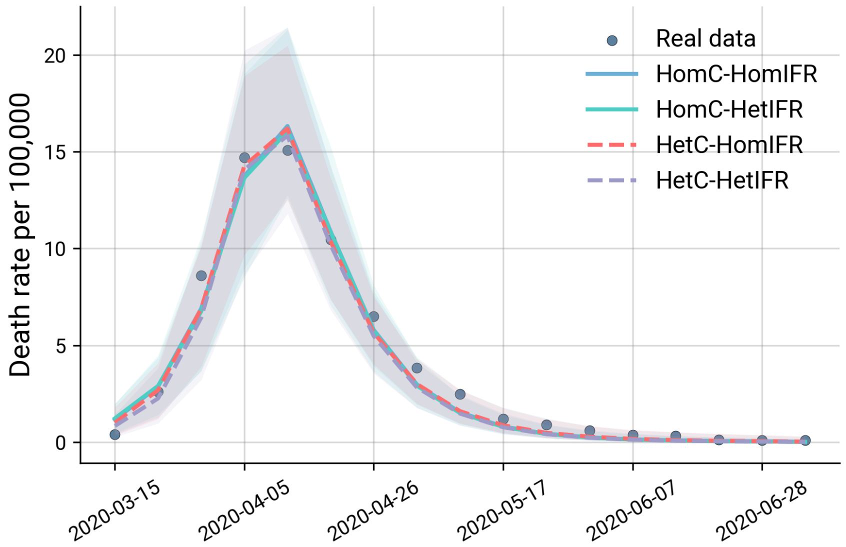

**Fig S3. Comparison of model fit to COVID-19 deaths in London.**

**Table S3.** Median wMAPEs between simulated disaggregated weekly deaths with respect to the reported deaths across age groups in London

| HomC-HomIFR | HomC-HetIFR | HetC-HomIFR | HetC-HetIFR |
|---|---|---|---|
| 1.9 | **0.6** | 2.4 | 1.0 |

#### 2.2 Fitting of COVID-19 deaths in NYC

We fit the models to COVID-19 weekly deaths in New York City during the first wave, spanning the same duration, from 2020-03-15 to 2020-07-05. Fig. S5 presents the models' fits to the aggregated deaths. All eight models exhibit a good fit to the overall data with comparable performance. To have a quantitative comparison, we calculate the wMAPEs between the data and the models' outcomes. According to Table S4, the two baseline models (Baseline-HomIFR/Baseline-HetIFR) yield the first and second lowest wMAPE, indicating the best fit to the aggregated death trajectory. However, the results change when evaluating the models' performance in fitting group-level data (see Fig. S6 and Table S5). The two baseline models report the highest median wMAPEs, indicating the poorest performance in capturing subgroup-specific characteristics. The Suscept-HetIFR model shows the lowest wMAPE, followed closely by the Prop-HetIFR and Assort-HetIFR models. In general, while the baseline model produces the best fit in terms of the aggregated weekly deaths, it fails to capture the heterogeneity observed in each subgroup. Models with group-specific susceptibility and heterogeneous mixing show more accurate fits to disaggregated data compared to the two baseline models.

**Table S4.** wMAPEs of simulated weekly deaths of models with respect to reported deaths in NYC

| Baseline-HomIFR | Suscept-HomIFR | Prop-HomIFR | Assort-HomIFR |
|---|---|---|---|
| **0.098** | 0.115 | 0.126 | 0.115 |
| Baseline-HetIFR | Suscept-HetIFR | Prop-HetIFR | Assort-HetIFR |
| 0.100 | 0.140 | 0.140 | 0.130 |

### 3 Models' fairness evaluation

#### 3.1 Relative concentration index of ordered social groups

Hereby, we present the Relative concentration index (RCI) of the models' outcome in the case of ordered social groups. The RCI is used to measure the inequality in weekly deaths across age groups (ordered from

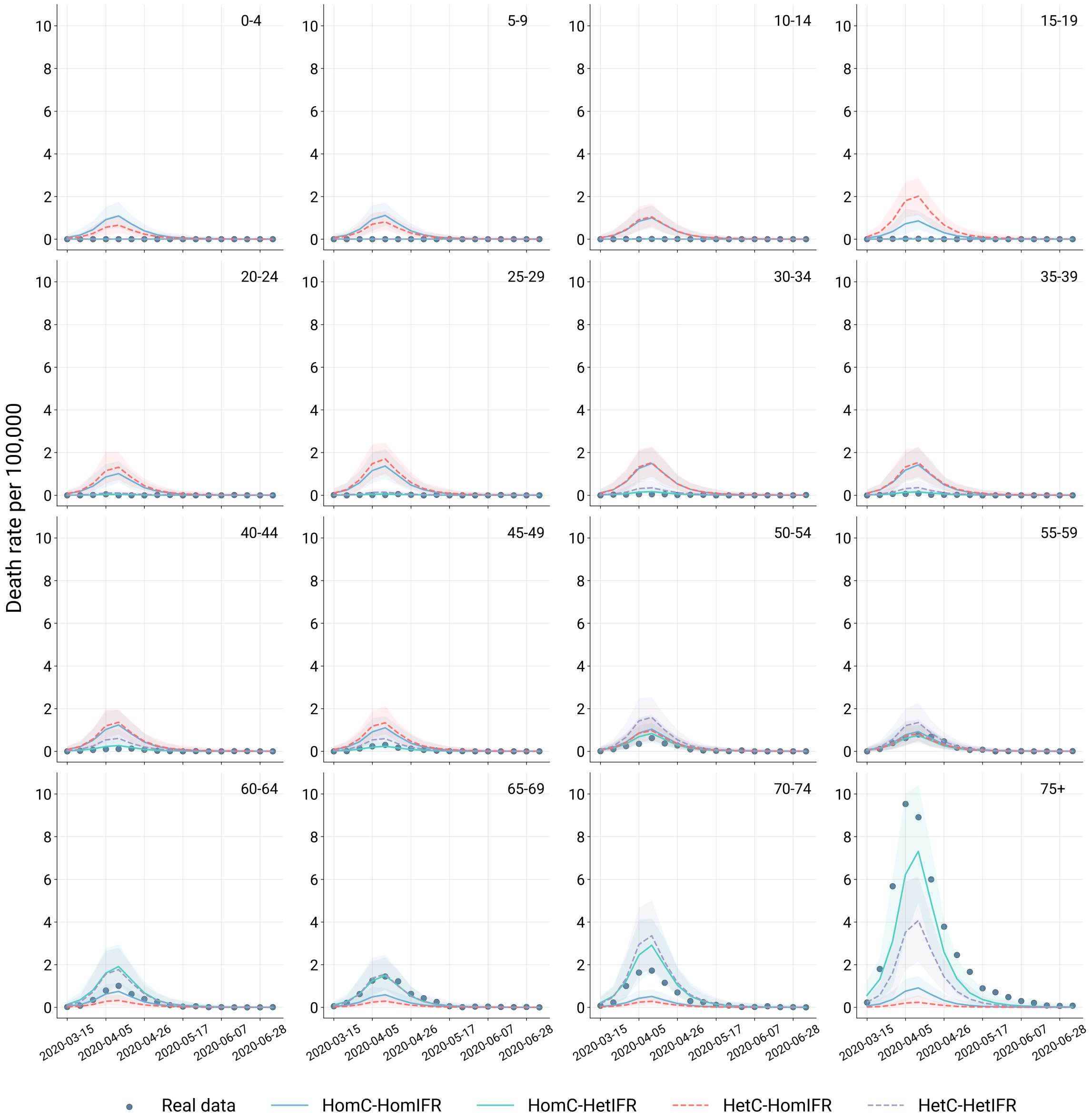

**Fig S4. Comparison of model fit to COVID-19 deaths by age in London.**

the youngest to eldest). RCI=0 indicates that death burdens are proportional to the population size of each age group. A negative RCI reflects disproportionate concentration of disease burdens among younger age groups, while positive RCI indicates a concentration among elder groups. Table S6 presents the RCI values of models' outcome and observed data. The real data reports a positive RCI of 0.8469, indicating the COVID-19 deaths are disproportionately concentrated among elder groups. The HomC-HetIFR model shows the closest RCI (0.8026) to the real data, and thereby reports the highest fairness score, followed by the HetC-HetIFR model. As expected, the HomoC-HomoIFR shows a value of RCI close to 0 as it assumes homogeneous contacts and IFR across age groups, leading to a uniform distribution of death burdens. The HetC-HomIFR model yields a negative RCI, suggesting a disproportionate burden of deaths among younger groups. Since the HetC-HomIFR model uses homogeneous IFR and the heterogeneous contact matrix where younger groups have intensive contacts than the elder.

**Table S5.** Median wMAPEs between simulated disaggregated weekly deaths with respect to the reported deaths across age groups in NYC

| Baseline-HomIFR | Suscept-HomIFR | Prop-HomIFR | Assort-HomIFR |
|---|---|---|---|
| 0.46 | 0.24 | 0.25 | 0.25 |
| Baseline-HetIFR | Suscept-HetIFR | Prop-HetIFR | Assort-HetIFR |
| 0.54 | **0.17** | 0.18 | 0.18 |

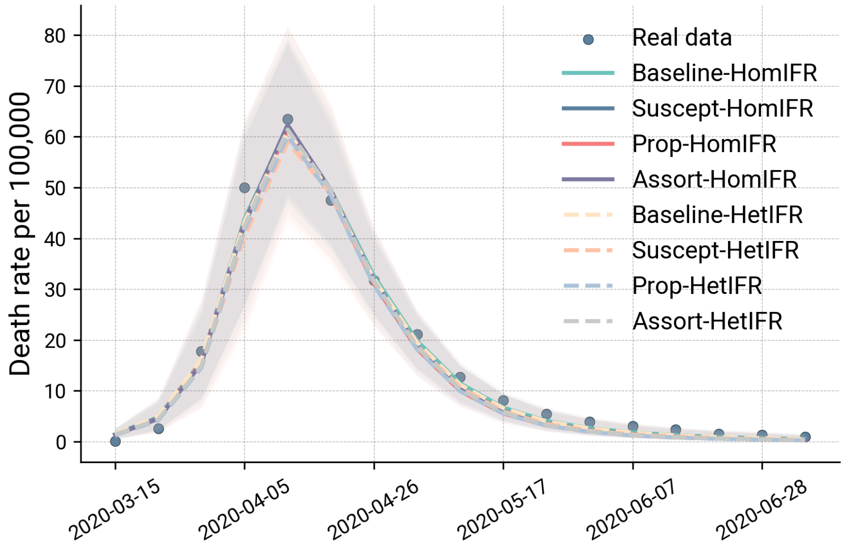

**Fig S5. Comparison of model fit to COVID-19 deaths in NYC.**

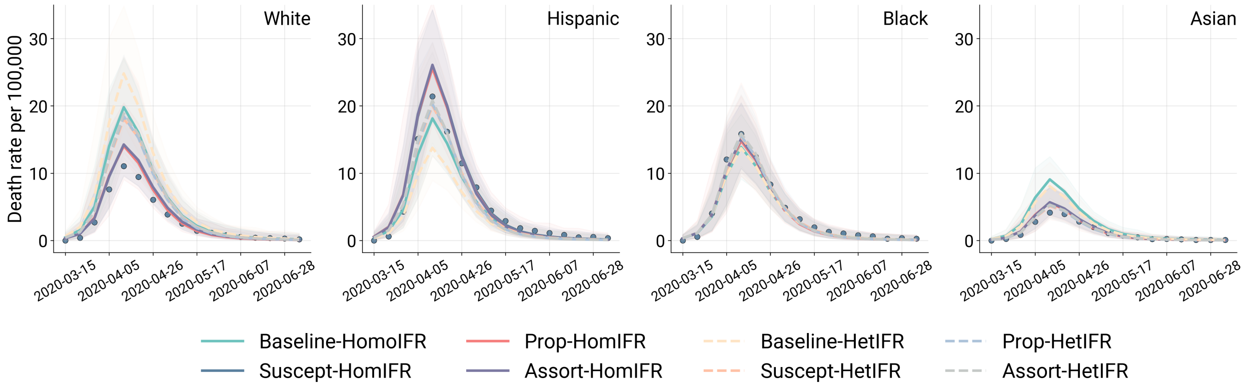

**Fig S6. Comparison of model fit to COVID-19 deaths by race and ethnicity in NYC.**

**Table S6.** RCI and fairness score of real deaths and the simulated results of models in London

| | RCI | Fairness score $F(age)$ |
|---|---|---|
| Surveillance data | 0.8469 | – |
| HomC - HomIFR | -0.0016 | 0.578 |
| HomC - HetIFR | 0.8026 | **0.978** |
| HetC - HomIFR | -0.0959 | 0.529 |
| HetC - HetIFR | 0.6967 | 0.925 |

#### 3.2 Theil index and fairness score in unordered social groups

We use the Theil index to quantify the inequality in COVID-19 deaths across racial/ethnic groups in NYC. A Theil index equal to 0 indicates that the disease burdens are proportional to the population size of each

**Table S7.** Theil index and fairness score of real deaths and the simulated results of models in NYC.

| | Theil index | Fairness score $F(race)$ |
|---|---|---|
| Surveillance data | 0.063 | – |
| Baseline-HomIFR | 1.78e-07 | 0.714 |
| Suscept-HomIFR | 0.0467 | 0.901 |
| Prop-HomIFR | 0.0470 | 0.901 |
| Assort-HomIFR | 0.0436 | **0.906** |
| Baseline-HetIFR | 0.0201 | 0.621 |
| Suscept-HetIFR | 0.0187 | 0.815 |
| Prop-HetIFR | 0.0186 | 0.815 |
| Assort-HetIFR | 0.0175 | 0.806 |

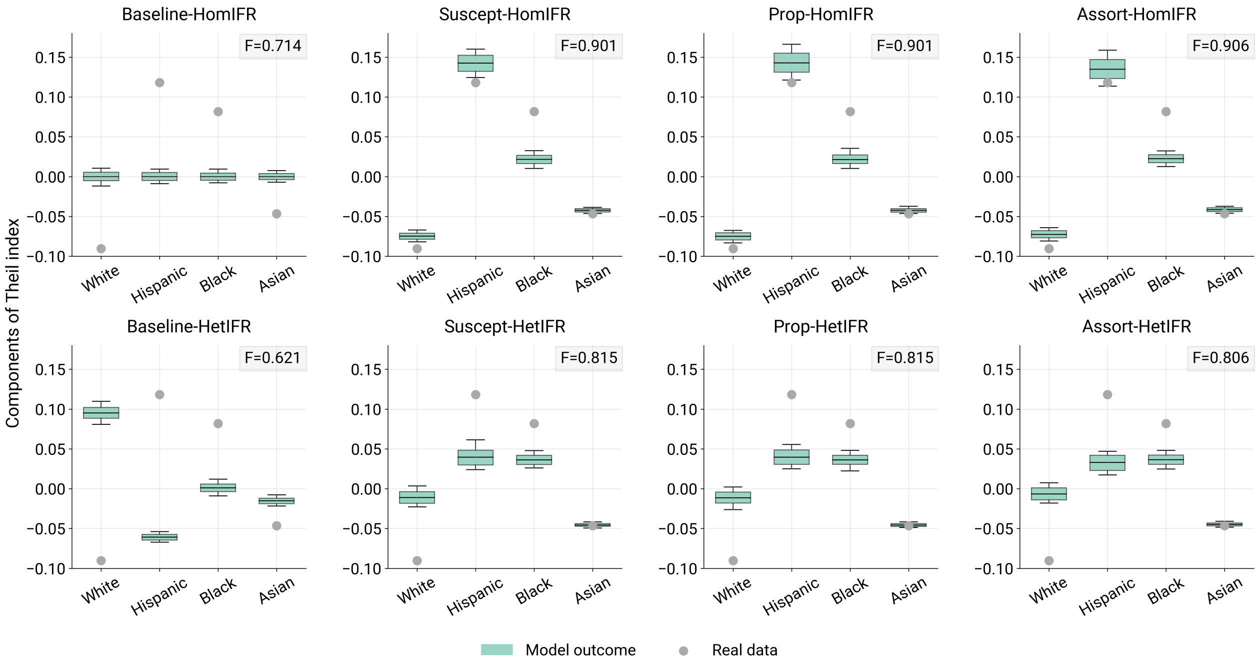

**Fig S7. Theil-index based fairness of all models considered to simulate the spread of COVID-19 in NYC.**

group. Larger values suggest greater inequality in the disease outcomes. As shown in Table S7, the Theil index of the data is 0.063. The Assort-HomIFR model yields the best fairness score among the eight models, indicating the best performance in capturing the empirical inequality patterns. Notably, the models with the closest Theil index to that of the data may not have the highest fairness score, since the fairness score is computed based on the components of Theil index for each group instead of the whole Theil index. In contrast, the Baseline-HetIFR model reports the worst score. The Baseline-HomIFR model, which assumes complete homogeneity in model parameters, shows near-zero values for each Theil index component. Besides, the models with heterogeneous IFR (HetIFR) consistently show worse performance than their counterparts with homogeneous IFR. This suggests that the age-adjusted IFRs may not accurately reflect real-world differences in COVID-19 IFR across racial or ethnic groups. Additionally, we compute and plot the Theil index components for each group to provide a clearer picture of how well the models fit each group (see Fig. S7.)

#### 3.3 Fairness of spatial models: the 2016 ZIKV epidemic in Colombia

As a second example of a spatially structured model, we examine the 2015-2016 Zika virus (ZIKV) outbreak in Colombia. Also in this case, we investigate the role of human mobility in capturing the heterogeneous burden of infections in space and compare the fairness of two modeling approaches that integrate different

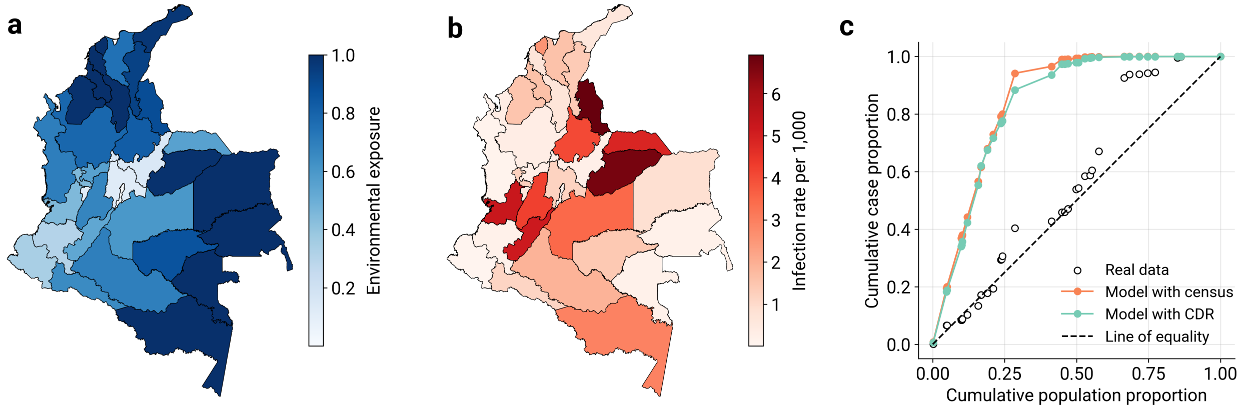

**Fig S8. Fairness and the 2016 Zika epidemic in Colombia (spatially structured models). a**, Choropleth map of Colombia, where departments are color coded according to the level of environmental exposure to the Aedes mosquitoes. **b**, ZIKV infection rates reported in each Colombian department, during the 2016 Zika epidemic. **c**, Comparison of the relative concentration curves of ZIKV cumulative incidence as reported by the surveillance (empty circles) and generated by two models: a spatial model with census mobility, and a model with mobile-phone derived mobility flows.

mobility networks. To this end, we consider the framework proposed by Perrotta et al. [15], who modeled the spread of ZIKV in Colombia with a model that incorporates human movements, either through mobile phone inferred mobility or through synthetic networks (see the Methods section for more details). ZIKV infection is a vector-borne disease transmitted primarily by infected *Aedes* mosquitoes; the risk of sustained disease transmission is highly dependent on local environmental conditions that can support the presence of the vector. Fig. S8a shows the spatial distribution of the environmental exposure index (EEI), a metric that captures the risk of exposure to the vector due to climatic and socioeconomic factors [15]. Fig. S8b shows the observed spatial heterogeneity of ZIKV incidence in Colombia in 2016, at the departmental level. If we consider the departments of Colombia, which are the main spatial units of analysis, as an ordered grouping based on environmental exposure, we can use the RCI to measure the inequality observed in the data and the simulation results. As shown in Fig. S8c, mobility-based epidemic models can reproduce the RCI curve of ZIKV infections only to a certain extent. However, it appears that the integration of mobility patterns derived from Call Detail Records (CDRs) slightly improves the fairness of the model ($F(\text{EEI}) = 0.74$), compared to a model that considers mobility flows as reported by the census ($F(\text{EEI}) = 0.73$). As we did for the case of Santiago, we can also consider the Colombian departments as unordered groups and compute the Theil index of ZIKV infections for the real data, the model with CDRs, and the model with census mobility. The fairness score based on the Theil index increases from $F = 0.205$ to $F = 0.215$, including mobility patterns from mobile phone data. We conclude that, in this epidemic scenario, an SLIR model with mobility derived from mobile phones can better capture the unequal spatial distribution of disease, whether we consider the population ranked by risk of environmental exposure or not.

### 4 Fairness of Interventions

#### 4.1 Vaccine allocation strategy

To show how the definition of models' fairness could contribute to guiding public health policies, taking the unordered social group case study in New York City as an example, we run a counterfactual vaccine intervention scenario and check the effects of different allocation strategies. For simplicity, we assume that the vaccination is implemented on July 6, 2020, hence right after the first wave of infections. We fully acknowledge that this scenario is not in line with the actual availability of COVID-19 vaccines outside of Mainland China. Hence, the analysis is purely illustrative. Furthermore, we simulate a second wave of infections by assuming the arrival of a new variant and a relaxation of NPIs (i.e., an increase in mobility levels). In particular, we continue running the simulations using the posteriors obtained by fitting the first epidemic wave as parameters. Also, we assume the emergence of a new variant with a 1.5 times higher transmission rate starting from 2020-07-06. From this date, we also assume flat, but higher mobility levels with respect to the last day of the first wave. This leads to a 1.4 times higher contact intensity among the population. The Theil-index based allocation strategy is designed based on the disease burden observed during the first wave.

We show the share of the number of vaccines distributed to each racial/ethnic group according to different vaccine strategies in Table S8. Based on the population based strategy, the number of vaccines allocated to the groups is proportional to the population size of the groups, where White has the largest share while Asian has the least vaccines. In the Theil-index based strategy, the Hispanic group gets the largest share of vaccines as they are the most vulnerable group and the second largest population group. In contrast, the Asian group receives the fewest vaccines, the share of which is even smaller compared to the population based, as it saw the fewest deaths in the first wave and is also the smallest group. In the combined strategy with $\gamma$=0.5, the White group receives a larger share of vaccines compared to the full Theil-index-based strategy, as more weight is placed on population in this case.

| **Strategy** | **Share of vaccines** | | | |
|---|---|---|---|---|
| | White | Hispanic | Black | Asian |
| Population based | 0.327 | 0.297 | 0.227 | 0.149 |
| Theil-index based | 0.227 | 0.421 | 0.232 | 0.119 |
| Combined ($\gamma = 0.5$) | 0.275 | 0.361 | 0.230 | 0.134 |

**Table S8.** Share of vaccines distributed to racial/ethnic groups according to different vaccine strategies. The sum of the share across the groups equal to 1 on each row.

#### 4.2 Sensitivity analysis on the number of vaccines

In the main text, we show the relative reduction in deaths given a vaccine coverage of $V_t$=20%. For sensitivity checks on the value of $V_T$, we explored the results obtained considering 10% and 30%. The results are shown in Table S9 and S10. Together with the results in the main text, these findings consistently show that the Theil-index-based strategy averts the largest number of overall deaths, though for $V_T$=30% the differences across allocation strategies are small, as the vaccination largely prevents a second outbreak (see Fig. S10). The weekly death trajectories for the four vaccine allocation strategies, given $V_T$=10% or 30%, are shown in Fig. S9 and S10.

| **Method** | **Relative reduction in deaths (%)** | | | | |
|---|---|---|---|---|---|
| | Overall | White | Hispanic | Black | Asian |
| Population based | 32.6 [25.7, 41.2] | **33.2** [26.1, 41.6] | 33.0 [25.6, 42.7] | 32.1 [24.8, 41.2] | **33.9 [26.4, 42.7]** |
| Theil-index based | **34.4** [26.7, 44.1] | 30.1 [22.1, 39.8] | **42.3** [33.8, 52.6] | **33.8 [26.0, 43.7]** | 32.9 [24.6, 42.5] |
| Combined ($\gamma = 0.5$) | 33.6 [26.3, 42.8] | 31.6 [24.4, 40.6] | 38.0 [30.1, 48.4] | 33.0 [25.4, 42.7] | 33.3 [25.3, 42.3] |

**Table S9.** Comparison of relative reduction in the number of deaths, by vaccine allocation strategy and racial/ethnic groups, given the number of vaccines $V_T$=10% of the population.

| **Method** | **Relative reduction in deaths** | | | | |
|---|---|---|---|---|---|
| | Overall | White | Hispanic | Black | Asian |
| Population based | 98.9 [82.8, 99.8] | 99.0 [83.3, 99.8] | 99.0 [84.1, 99.8] | 98.9 [82.4, 99.9] | 99.0 [84.2, 99.9] |
| Theil-index based | **99.3** [86.2, 99.8] | 99.1 [82.0, 99.8] | **99.6** [93.7, 99.9] | **99.3 [86.2, 99.9]** | **99.3 [85.4, 99.9]** |
| Combined ($\gamma = 0.5$) | 99.2 [85.3, 99.8] | **99.1** [83.1, 99.8] | 99.4 [90.0, 99.9] | 99.2 [85.2, 99.9] | 99.2 [85.3, 99.9] |

**Table S10.** Comparison of relative reduction in the number of deaths, by vaccine allocation strategy and racial/ethnic groups, given the number of vaccines $V_T$=30% of the population.

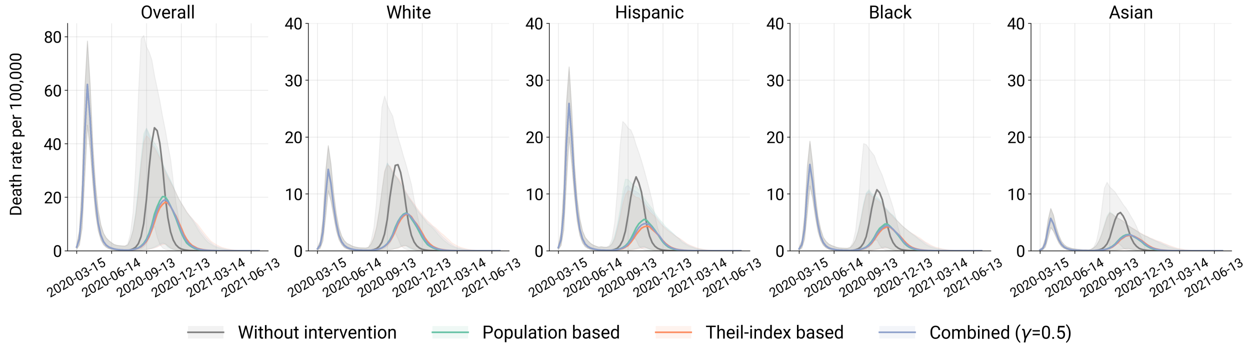

**Fig S9. Comparison of vaccine allocation strategies on weekly deaths by race/ethnicity in NYC, given $V_T$=10% of the population.**

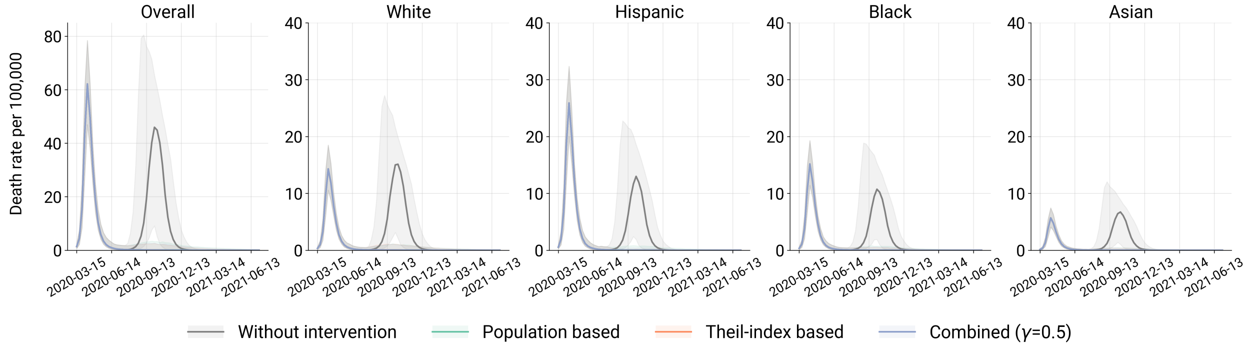

**Fig S10. Comparison of vaccine allocation strategies on weekly deaths by race/ethnicity in NYC, given $V_T$=30% of the population.**

[4] Google llc "google covid-19 community mobility reports". `DOI:https://www.google.com/covid19/mobility/`.
